# Olfactory cleft obstruction in post-COVID-19 olfactory disorder: CT Comparison with post-viral cases

**DOI:** 10.1101/2025.06.04.25328964

**Authors:** Hirotaka Tanaka, Eri Mori, Yuji Kishimoto, Nagomi Yonezawa, Rumi Sekine, Monami Nagai, Masayoshi Tei, Nobuyoshi Otori

## Abstract

**Background:** Post-COVID-19 olfactory dysfunction (PCOD) is a common sequela of SARS-CoV-2 infection, with some cases persisting beyond the acute phase. While prolonged PCOD has been considered primarily sensorineural, recent studies suggest that olfactory cleft (OC) obstruction may contribute to olfactory dysfunction (OD). This study aimed to investigate OC obstruction in prolonged PCOD compared to post-infectious olfactory dysfunction (PIOD) and assess its impact on olfactory function.

**Methodology/Principal:** We retrospectively analysed sinus computed tomography scans of patients with prolonged PCOD and PIOD. OC obstruction was classified into absent (0%), mild (1–90%), and severe (>90%) types. The severity of OC obstruction was compared between PCOD and PIOD cases, and olfactory test results were analysed according to OC obstruction severity.

**Results:** A total of 87 PCOD patients and 67 PIOD patients were included. Mild and severe OC obstruction was more prevalent and severe in prolonged PCOD (mild: 35.6%, severe: 18.4%) than PIOD (mild: 13.4%, severe: 3%). Moreover, worsened olfactory function was found in PCOD patients with severe OC obstruction. In contrast, OC obstruction showed little association with olfactory function in PIOD patients.

**Conclusions:** Mild and Severe OC obstruction appear to be more common in prolonged PCOD. Moreover, severe OC obstruction may further impair olfactory function in PCOD patients. Conversely, severe OD in PIOD *patients occurred independently of OC obstruction*.

## INTRODUCTION

Approximately half of coronavirus disease 2019 (COVID-19) cases develop post-COVID-19 olfactory dysfunction (PCOD) ^(1)^, which typically resolves within weeks to months following infection ^(2)^. Eliezer et al. reported that complete obstruction of the olfactory cleft (OC) was observed in 95% of acute PCOD patients, with 63% demonstrating resolution of both OC obstruction and olfactory dysfunction (OD) within one month ^(3)^. Therefore, acute PCOD accompanied by OC obstruction is expected to be due to conductive rather than sensorineural OD.

Conversely, residual PCOD persists in approximately 8–20% of patients even after one year ^(4,5)^. PCOD has been classified as sensorineural OD ^(6)^, similar to non-COVID-19-associated post-infectious OD (PIOD). However, Kandemirli et al. reported that OC opacification was present in 73.9% of prolonged PCOD cases ^(7)^. Furthermore, a systematic review and meta-analysis of radiological imaging of PCOD found that OC opacification was prevalent in 27.5–63% of PCOD cases, suggesting a contributory role of conductive OD ^(8,9)^. These findings indicate that a subset of persistent PCOD cases may involve OC obstruction as a potential factor in OD pathogenesis.

Although several reports have described OC opacification as a characteristic finding of PCOD, it remains unclear whether this radiological feature is also common in PIOD. A recent magnetic resonance imaging (MRI)-based study by Li et al. ^(10)^ demonstrated that lower OC opacification scores in PIOD patients compared to PCOD patients, suggesting possible differences in underlying pathophysiology. However, MRI has limitations in spatial resolution particularly for visualising the fine structures of the OC. In contrast, paranasal sinus CT offers superior spatial resolution and is well-suited for assessing detailed anatomical findings in the OC. To date, no studies have systematically compared OC obstruction between PCOD and PIOC using CT beyond the acute phase.

In this study, we aimed to compare OC obstruction on coronal sinus CT scans with PCOD and PIOD. Additionally, we examined whether the severity of OC obstruction is associated with olfactory function in PCOD and PIOD, respectively.

## MATERIALS AND METHODS

### Study design and participants

This study was approved by the ethics committee of Jikei University School of Medicine (Approval No. 33-159[10774]). We included patients diagnosed with PIOD who visited the Department of Otorhinolaryngology, Jikei University Hospital between 1 January 2018, and 31 December 2019, as well as those diagnosed with PCOD (based on positive PCR tests at the time of onset of OD) who visited between 1 January 2020, and 31 May 2024. In Japan, the first confirmed case of SARS-CoV-2 infection was reported on 15 January 2020; therefore, we assumed that PIOD cases prior to this date were not influenced by COVID-19. The diagnosis of OD was made comprehensively by olfactory specialists based on medical interviews, nasal endoscopy, imaging studies (such as sinus CT and MRI), and olfactory tests, in accordance with the Japanese Clinical Practice Guideline for Olfactory Dysfunction ^(11)^.

### Variables

We analysed clinical information and olfactory test results in patients with PCOD and PIOD. Collected clinical data included age, sex, medical history, family history, smoking status, duration of OD, CT findings, and olfactory test results. Symptoms including parosmia, or distorted sense of smell, and phantosmia, or perception of smell in the absence of an odourant, were also collected. Regarding smoking status, patients were categorised as current, or former or never-smokers, based on previous reports suggesting that current smokers have an increased risk of developing OD ^(12,13)^.

### OC assessment by coronal sinus CT scan

CT examinations were performed using a 128 × 2 detector dual-source CT scanner (SOMATOM Drive; Siemens, Erlangen, Germany). In this study, we focused on the region between the superior turbinate and nasal septum within the OC. The degree of obstruction was comprehensively evaluated across all coronal slices from the anterior to posterior end of the superior turbinate.

Three otorhinolaryngologists evaluated the coronal CT images. To minimize inter-rater variability and ensure the assessment reliability, all raters underwent standardised training prior to the evaluation process. Specifically, the initial 30 cases were reviewed jointly through group discussion to align the evaluation criteria among the three raters. The remaining cases were assessed independently. Following individual assessments, results were compared, and any discrepancies were resolved through consensus discussions. The following classification criteria were used for the evaluation of OC obstruction (Figure 1):

**Figure 1.**
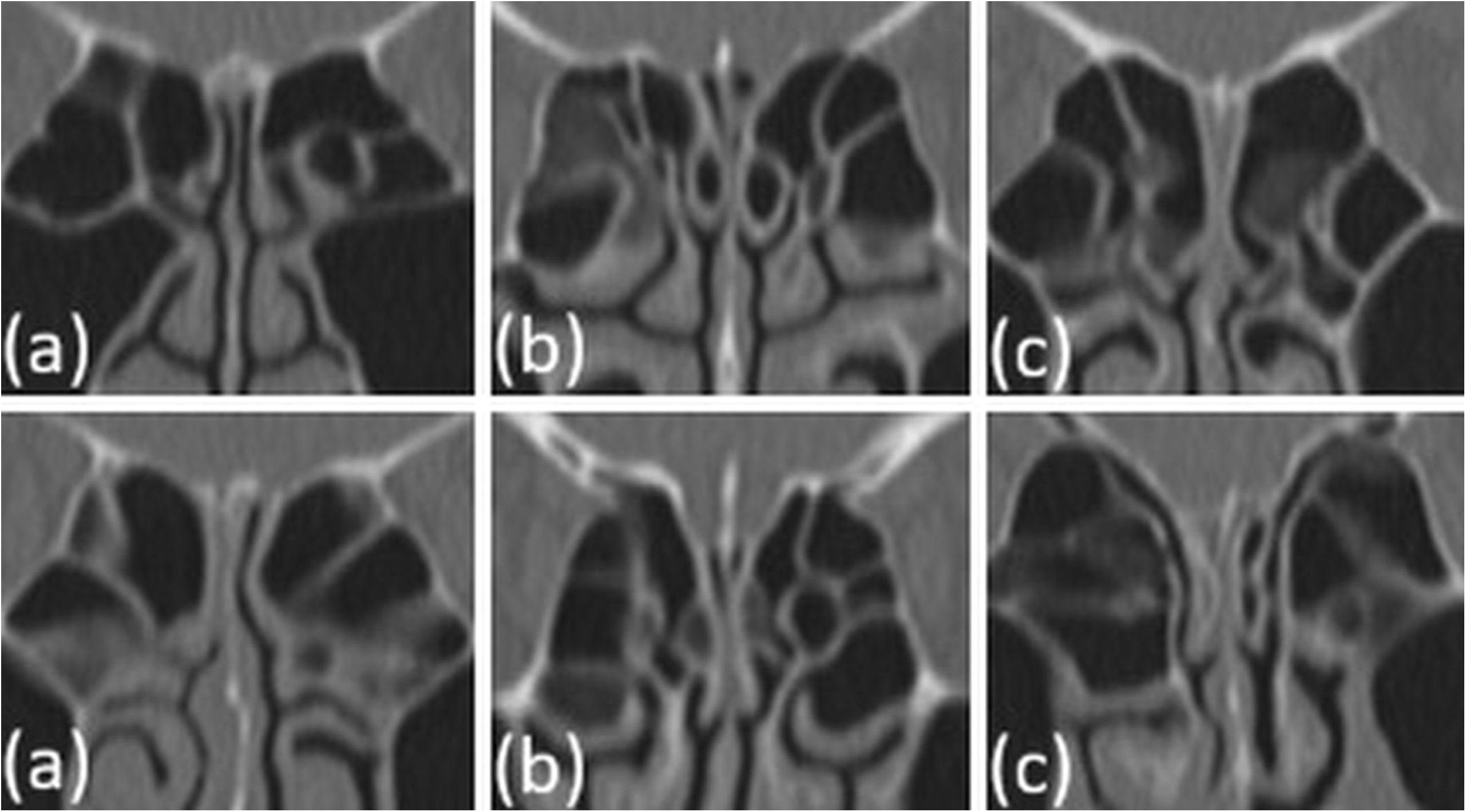
Classification of bilateral OC obstruction on coronal sinus CT. (a) Absent: No contact between the superior turbinate and nasal septum on at least one side of the OC. (b) Mild: Bilateral contact present but obstruction is less than 90% on at least one side. (c) Severe: Extensive bilateral contact with more than 90% obstruction on both sides. OC, olfactory cleft; CT, computed tomography.

- Severe: Extensive bilateral contact between the superior turbinate and nasal septum, obstructing more than 90% of the surface on both sides.
- Mild: Bilateral contact present, but with less than 90% obstruction on at least one side.
- Absent: No contact on at least one side of OC.

### Olfactory tests

The following olfactory tests were performed:

- Sensory tests

⟶ T&T olfactometer test (T&T): Standard olfactory test widely used in Japan, employing five olfactory substances: β-phenylethil alcohol, methyl cyclopentenolone, isovaleric acid, γ-undecalactone, and skatole. Eight concentration levels, ranging from minus 2 to 5, were prepared in 10-fold serial dilutions, with level 0 representing the olfactory threshold of healthy adults. Patients sniffed each odour starting from the lowest concentration (minus 2). The concentration at which the odour was first detected was recorded as the detection threshold. The concentration at which the patient could identify the smell was recorded as the recognition threshold. If the patient failed to detect the odour, a score of 6 was assigned (expect for methyl cyclopentenolone, which was assigned a score of 5). The average detection and recognition thresholds were calculated by dividing the total score by five. Based on guideline criteria ^(14)^, patients were classified as follows:

- Normosmia: Recognition threshold, < 1.1
- Mild olfactory dysfunction: ≤ 2.5
- Moderate olfactory dysfunction: ≤ 4
- Severe olfactory dysfunction: ≤ 5.5
- Anosmia ≥ 5.6
⟶ Open Essence (OE): Card-type odour identification test consisting of 12 folded cards, each releasing a specific odour when opened. The tested odourants included calligraphy ink, wood, perfume, menthol, orange, curry, cooking gas, rose, cypress wood (Japanese cypress, ‘hinoki’), sweaty clothes, condensed milk, and roasted garlic. Patients were instructed to select an answer from six choices, which included ‘cannot identify’, ‘odourless’, and four odour names (one correct answer and three distractors) ^(15)^. Patients were classified as normosmic if they correctly identified ≥8 of the 12 odours, whereas those with ≤7 correct answers were classified as having OD ^(16)^.
- Questionnaires

⟶ Visual analogue scale for OD (VAS): A 100-mm straight line was used to assess olfactory function, with the left end representing ‘cannot smell at all’ (0 points) and the right end representing ‘normal olfaction’ (100 points). Patients marked their current odour perception along this line, with the score being the distance (in millimetres) from the left edge to the marked point.

⟶ Self-administered odour questionnaire (SAOQ): Participants were asked to rate how well they were able to smell 20 odours commonly encountered in daily life in Japan, using a four-point scale with the following points assigned: ‘always smelled’ (2 points), ‘sometimes smelled’ (1 point), ‘never smelled’ (0 points), and ‘unknown or no recent exposure (to odourant)’. Items rated as ‘unknown or no recent exposure’ were excluded from the final evaluations. The total score was converted into a percentage for assessment ^(17)^.
Retronasal olfactory test

⟶ Intravenous olfactory test, or Alinamin Test (AT): Subjective retronasal olfactory test widely used in Japan, which utilises the property of Alinamin^®^, that one of its degradation products, propyl mercaptan, has a garlic-or onion-like odour. Alinamin^®^ (prosultiamine, 10 mg, 2 mL; Takeda Pharmaceutical Company) was injected into the medial cubital vein at a constant rate for 20 seconds. After circulating in the bloodstream, its degradation products diffused into the lungs and were then exhaled. Patients were instructed to take nasal breaths quietly every two seconds. When these compounds reached the olfactory epithelium through the posterior choana, they stimulated olfactory neurons, resulting in the perception of the garlic-or onion-like odour. Two parameters were evaluated: latency time, which was the time from the start of injection to odour perception; and duration time, which was the time from odour perception until disappearance. If the patient was unable to detect any odour, it was recorded as ‘no response’, which indicates a poor prognosis for OD ^(18)^. Latency time > 8 seconds was categorised as ‘prolonged latency’, and duration time < 70 seconds was categorised as ‘shortened duration’ ^(14)^.

### Exclusion criteria

Patients were excluded from the study if they met any of the following criteria:

- Normosmia on the T&T test at the first visit.
- Symptom of unilateral OD.
- Disease duration of less than 2 months before the first visit (to exclude the acute phase).
- Did not undergo a sinus CT scan at the first visit.
- Sinus CT scan revealed soft tissue opacities in the ethmoidal sinus (Lund-Mackay score of 1 or higher).
- History of sinus surgery or skull base surgery.

### Outcomes

The primary outcome was to compare OC obstruction between PCOD and PIOD patients. The secondary outcome was to investigate the relationship between the severity of OC obstruction and the results of olfactory tests in PCOD or PIOD patients.

### Statistical analysis

Continuous variables are presented as medians [interquartile ranges (IQRs)], while categorical variables are expressed as counts (%). Comparison between PCOD and PIOD patients were performed using Fisher’s exact test for categorical variables: sex, age, disease duration, parosmia, phantosmia, smoking status, and OC obstruction by sinus CT scan. The Mann–Whitney U test was used for continuous variables: detection and recognition thresholds of T&T, OE, VAS, SAOQ, and AT results.

Comparisons among the three severity levels of OC obstruction were conducted using Fisher’s exact test for categorical variables: sex, age, disease duration, parosmia, phantosmia, and smoking status. The Kruskal-Wallis test was used for continuous valuables: detection and recognition thresholds of T&T, OE, VAS, SAOQ, and AT results. The Dunn-Bonferroni post-hoc test was used to adjust the significance level for multiple comparison.

A two-sided p value of < 0.05 was considered statistically significant. Statistical analyses were performed using GraphPad Prism version 8.4.3 (GraphPad Software Inc., San Diego, CA, USA), unless otherwise indicated.

## RESULTS

### Background of patients

A total of 230 patients visited our outpatient department during the study period, including 127 with PCOD and 103 with PIOD. Of these, 87 PCOD patients and 67 PIOD patients were included in this study according to the exclusion criteria (Figure 2). Table 1 presents the background characteristics of both groups. The median age of PCOD patients was 41 years (range, 11–81), which was significantly younger than PIOD patients (63 years, range, 18–88) (p < 0.000). A significantly higher proportion of males was observed in the PCOD group compared to the PIOD group (41.3% vs 19.4%, p = 0.005). Median duration of OD was significantly longer in PCOD patients (6.0 months vs 5.0 months, p = 0.038). Parosmia was also significantly more prevalent in PCOD patients (29.8% vs 11.9%, p = 0.010). No significant differences were found between the two groups in smoking status or phantosmia.

**Figure 2.**
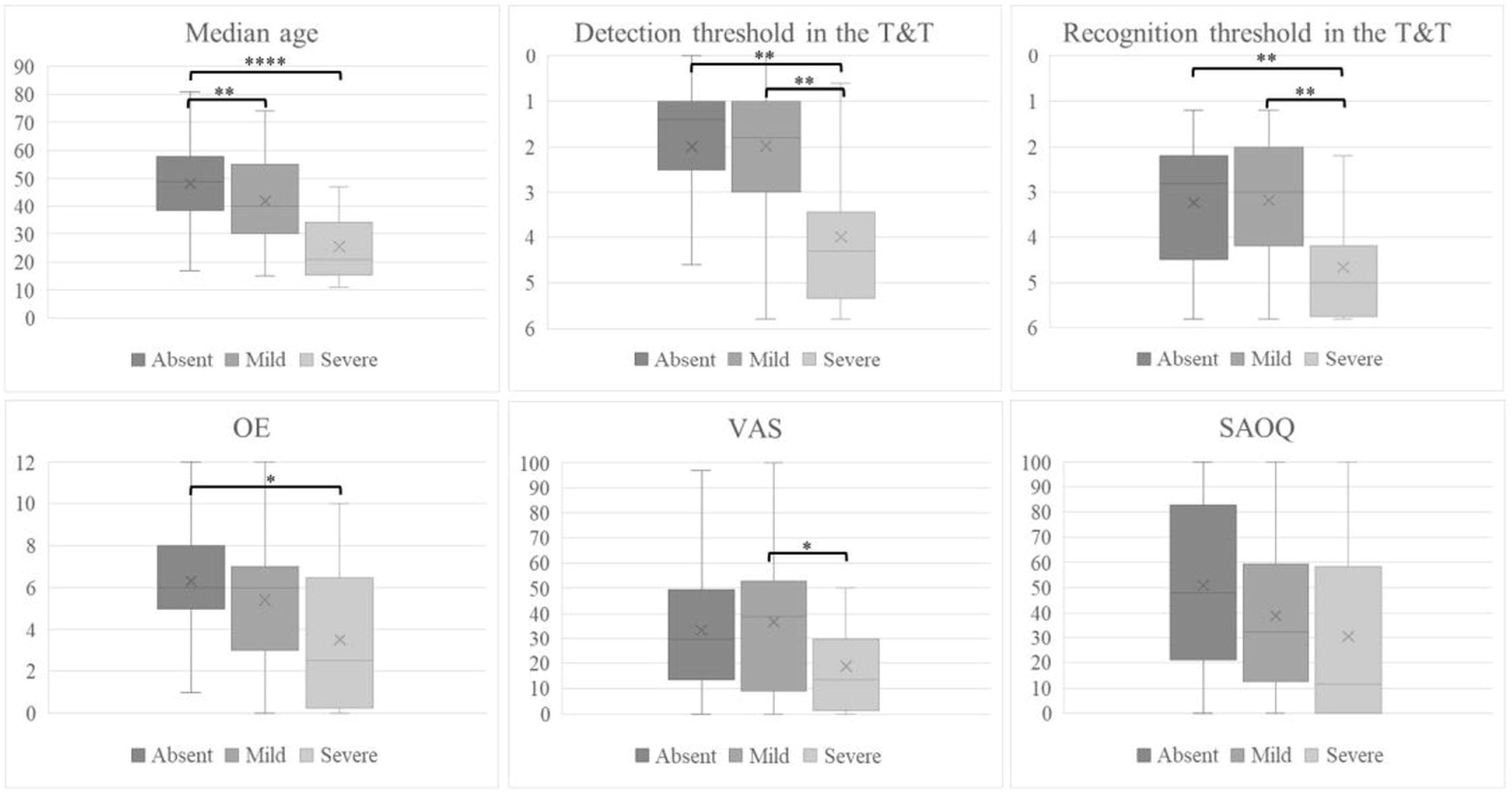
Boxplot of median age and olfactory test results by OC obstruction severity in PCOD patients. Boxplot illustrating the distribution of median age and olfactory test scores showing significant differences across different severities of OC obstruction in PCOD patients. Pairwise comparisons were performed using the Kruskal-Wallis test, with p-values adjusted using the Dunn-Bonferroni post-hoc test for multiple comparisons. OC, olfactory cleft; PCOD, post-COVID-19 olfactory dysfunction; T&T, T&T olfactometer test; OE, Open Essence; VAS, visual analogue scale for olfactory dysfunction; SAOQ, self-administered odour questionnaire. * p < 0.05, ** p < 0.01, **** p < 0.000

### Comparison of olfactory tests and OC obstruction between PCOD and PIOD (Table 1)

In olfactory tests, PCOD patients showed significantly better results compared to PIOD patients across almost all measures:

– Detection and recognition thresholds of the T&T: 1.8 vs 4.4, and 3.2 vs 5.4, respectively, both p < 0.000
– VAS: 30.0 vs 8.0 points, p < 0.000
– SAOQ: 41.7% vs 10.3%, p = 0.002
– ‘No response’ to AT: 11.4% vs 43.5%, p < 0.000

Regarding OC obstruction on sinus CT scans, PCOD patients had a significantly higher prevalence of combined severe and mild OC obstruction than PIOD (54.0% vs 16.4%, p < 0.000). Additionally, while only 2 cases of severe OC obstruction (3.0%) were observed in PIOD patients, the number was significantly higher in PCOD patients (16 cases, 18.4%, p = 0.004).

### Relationship between OC obstruction and olfaction in PCOD and PIOD patients (Table 2 and Table 3)

Table 2 and 3 compare the background characteristics and olfactory test results of PCOD and PIOD patients, categorised into three groups based on the severity of OC obstruction.

Among PCOD patients (Table 2), there were no significant differences in sex, disease duration, parosmia, phantosmia, or smoking status across the groups. The median age was significantly younger in patients with severe OC obstruction than in those with absent (p < 0.000) or mild OC obstruction (p = 0.007). In the olfactory tests, detection and recognition thresholds of the T&T were significantly worse in patients with severe OC obstruction than in those with absent or mild OC obstruction (p = 0.001 and 0.002, p = 0.003 and 0.003, respectively). OE scores were significantly worse in patients with severe OC obstruction than in those with absent obstruction (p = 0.012). VAS scores were significantly worse in patients with severe OC obstruction compared to those with mild obstruction (p = 0.042). No significant differences were found in SAOQ and AT results among the three groups (Figure 2).

In contrast, among PIOD patients (Table 3), 9 cases (13.4%) had mild OC obstruction, and 2 cases (3.0%) had severe OC obstruction. No significant differences in background characteristics or olfactory test results were observed across the groups.

## DISCUSSION

Our study revealed three key findings regarding OC obstruction in prolonged PCOD and PIOD. First, OC obstruction—particularly severe bilateral obstruction—was significantly more prevalent in the PCOD group compared to the PIOD group. Second, among PCOD patients, severe OC obstruction was more frequently observed in younger individuals and was associated with worse OD. Third, in the PIOD group, OD appeared to be independent of the presence or severity of OC obstruction.

### Conductive OD may be a hidden contributor of prolonged PCOD

SARS-CoV-2 infection of the olfactory mucosa is mediated by angiotensin-converting enzyme (ACE2) and type II transmembrane serine protease (TMPRSS2), both predominantly expressed in sustentacular cells and Bowman’s glands rather than in olfactory receptor neurons ^(19-21)^. This viral infection induces inflammation in these cells, leading to mucosal swelling and increased mucus production in the OC, ultimately resulting in conductive OD. Typically, as the infection resolves, the mucosal swelling subsides and OD improves rapidly ^(3)^. However, in some patients, PCOD persists, likely due to viral damage to olfactory receptor neurons, resulting in a pathology similar to conventional PIOD ^(4)^.

Although, our study found that the OC obstruction was more frequent in PCOD than in PIOD, OD appeared milder in PCOD group. This discrepancy might suggest a limited impact of OC obstruction on olfactory function. However, bilateral OC obstruction was observed in approximately half of the PCOD patients beyond the acute phase - a rate more than three times higher than in PIOD. Notably, severe bilateral OC obstruction was found in 18% of PCOD patients, which was sixfold higher than in PIOD. Furthermore, while no clear relationship between OC obstruction and olfactory function in the PIOD group, PCOD patients with absent or mild OC obstruction had relatively preserved olfactory function, whereas those with severe OC obstruction showed significantly worse OD. These findings suggest that, unlike PIOD, olfactory function in PCOD may be more susceptible to the degree of OC obstruction, potentially skewing overall olfactory scores toward a milder presentation. This could partially explain why PCOD appears milder overall, as the group includes many patients with minimal obstruction. Considering that OC obstruction can impair airflow and lead to conductive OD ^(22-25)^, these results further support the hypothesis that a conductive component may underlie prolonged OD in a subset of PCOD patients.

As for the correlation between OC obstruction and olfactory test results, no significant associations were found between the severity of OC obstruction and the latency and duration times, or absence of response to the AT. However, a closer look at individual PCOD cases revealed that 4 of 5 anosmic patients with severe OC obstruction responded to the AT (the remaining case was not tested), whereas only 1 of 7 anosmic patients with absent or mild OC obstruction showed a response. Pfaar et al. previously demonstrated that occluding the anterior portion of the OC with a sponge reduces orthonasal olfactory function while sparing retronasal olfaction ^(26)^. These findings highlight the value of distinguishing between orthonasal and retronasal olfaction, and performing olfactory testing for both. Even if orthonasal testing indicates anosmia, retronasal olfaction may be spared, suggesting an underlying conductive mechanism. These findings further support the notion that PCOD with severe OC obstruction may involve conductive OD. Based on these observations, we propose that prolonged PCOD may represent, at least in part, conductive OD attributable to OC obstruction.

### PCOD with severe OC obstruction may worsen olfactory function and may be more common in young people

Previous studies on the relationship between OC obstruction and OD have yielded conflicting results, with some reporting a correlation ^(27,28)^ and others finding no association ^(29)^. However, these investigations employed various methodologies, such as including unilateral OC obstruction, only examining presence or absence of OC obstruction and not its severity, and defining severe OC obstruction as contact exceeding two-thirds. Because olfactory tests typically require simultaneous sniffing from both nostrils, impaired olfactory function can only be detected if both sides are affected. Moreover, as demonstrated in our study, if OC obstruction is mild, airflow to the OC may be preserved, and it may not contribute to OD. Thus, the prevalence of OC obstruction in OD cases and its precise relationship with olfactory function remains unclear.

Interestingly, in this study, severe OC obstruction was more frequently observed in younger PCOD patients. In 1958, Takahashi et al. investigated cadaveric OC specimens and found synechiae or contact between the superior turbinate and nasal septum in 4 sides (8%) among 23 cases (46 sides) under 12 years of age, and 39 sides (48%) among 41 cases (82 sides) over 17 years of age ^(30)^. By contrast, Koo et al. examined sinus CT scans in individuals aged 17 years and older and reported that the prevalence of concha bullosa in middle and superior turbinates did not differ significantly across age groups ^(31)^. These findings suggests that morphological changes in the turbinate can occur during maxillofacial development from childhood to adolescence, potentially leading to narrowing of the OC. However, no comprehensive literature has systematically examined changes in sinus CT findings by age, and it remains unclear whether younger individuals physiologically exhibit a narrower or more obstructed OC.

### Inflammatory obstruction may decrease over time

Although disease duration of PCOD differed significantly among the absent, mild and severe OC obstruction groups, post hoc comparisons did not reveal statistically significant differences between individual groups. However, the median disease duration was longest in the absent group and longer in the mild OC obstruction group compared to the severe group. This trend suggests that the degree of OC obstruction may decrease over time following the acute phase.

Based on Jiang et al., by analysing CT findings related to OC obstruction in detail, OC obstruction was classified into the following three types ^(32)^:

- Structural obstruction – caused by contact between the nasal septum and turbinates, often due to concha bullosa or medial deviation (Figure 3a).
- Inflammatory obstruction – characterized by soft tissue opacification occupying the OC (Figure 3b).
- Mixed obstruction – involving both structural and inflammatory components (Figure 3c).

**Figure 3.**
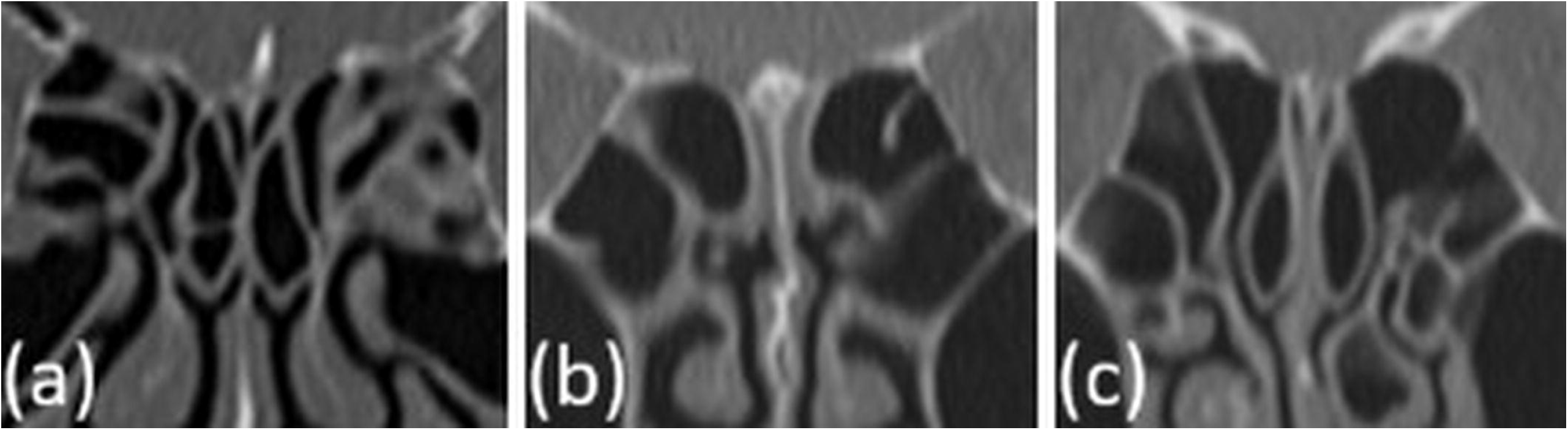
Type of OC obstruction based on coronal sinus CT findings. (a) Structural obstruction: Caused by contact between the nasal septum and turbinates due to concha bullosa or medial deviation. (b) Inflammatory obstruction: Characterized by soft tissue opacification filling the OC. (c) Mixed obstruction: Combination of both structural and inflammatory components. OC, olfactory cleft; CT, computed tomography.

A recent study on olfactory mucosa biopsies in long COVID patients indicated that inflammation could persist in the olfactory epithelium long after SARS-CoV-2 has cleared, suggesting a potential mechanism underlying prolonged PCOD ^(33)^. Moreover, it is unlikely that SARS-CoV-2 infection would newly induce structural abnormalities such as concha bullosa or medial deviation of the superior turbinate. Therefore, it is possible that the inflammatory OC obstruction decreased with time after the acute phase. To clarify how OC obstruction and olfactory function change over time, further longitudinal studies are warranted.

### Limitations

First, this was a retrospective study and it remains unclear whether sinus CT scans are necessary for all cases of PCOD and PIOD. According to the International Consensus Statement ^(34)^, “In case of suspected OC syndrome or sinonasal disease causing OD, CT scan can be considered as an option to confirm the diagnosis.” Thus, the indication for sinus CT in patients with prolonged PCOD or PIOD should be evaluated individually based on clinical presentation.

Second, the role of non-severe OC obstruction in OD remains undetermined.

Third, this study focused on OC obstruction between the superior turbinate and nasal septum, and obstruction between the middle turbinate and nasal septum was not assessed.

Nonetheless, we believe that this study is novel in highlighting OC obstruction as a potential contributor in prolonged PCOD. Furthermore, it underscores the importance of evaluating the OC via sinus CT scans to identify the underlying causes of persistent PCOD.

### Conclusion

Our findings suggest that, compared to PIOD, OC obstruction—particularly when severe—may be associated with the severity of OD in prolonged PCOD. Additionally, severe OC obstruction was more frequently observed in younger individuals and may further impair olfactory function in PCOD patients. In contrast, severe OD was observed regardless of the presence of OC obstruction in PIOD patients.

## Data Availability

All data produced in the present study are available upon reasonable request to the authors

## ACKNOWLEDGEMENTS

We would like to thank Dr. Miyuki Tanino for English language editing.

We would like to thank Professor Hiroya Ojiri at the Department of Radiology, Jikei University School of Medicine, for his valuable advice regarding the radiological evaluation of OC abnormalities.

## AUTHORSHIP CONTRIBUTION

Hirotaka Tanaka: conceptualization, data collection and analysis, funding acquisition, investigation, methodology, resources

Eri Mori: conceptualization, investigation, methodology, writing, review, and editing of the manuscript, and total project administration and supervision.

Yuji Kishimoto & Nagomi Yonezawa & Rumi Sekine & Monami Nagai: data collection and carrying out the olfactory tests

Masayoshi Tei: review and editing of the manuscript

Nobuyoshi Otori: supervision, review, and editing of the final manuscript.

