## Supplementary material for "Olfactory cleft obstruction in post-COVID-19 olfactory disorder: CT Comparison with post-viral cases": Tables

| Table 1. Demographic characteristics and olfactory test results of PCOD and PIOD patients | | | | | | | | |
| --- | --- | --- | --- | --- | --- | --- | --- | --- |
| Characteristics | | PCOD (n=87) | |  | PIOD (n=67) | |  | p |
| Sex, No (%) | |  |  |  |  |  |  |  |
|  | Male | 36 | (41.4) |  | 13 | (19.4) |  | 0.005* |
|  | Female | 51 | (58.6) |  | 54 | (80.6) |  |  |
| Age, median yr (IQR) | |  |  |  |  |  |  |  |
|  |  | 41.0 | (27.0-54.5) | | 63.0 | (51.5-70.0) | | <0.000**** |
| Disease duration, median month (IQR) | | |  |  |  |  |  |  |
|  |  | 6.0 | (4.0-11.0) |  | 5.0 | (3.0-7.0) |  | 0.038* |
| Smoking, No (%) | |  |  |  |  |  |  |  |
|  | Current | 4 | (5.7) |  | 5 | (7.5) |  | 0.493 |
|  | Former or never | 83 | (94.3) |  | 58 | (86.6) |  |  |
|  | Unknown | 0 | (0) |  | 4 | (6.0) |  |  |
| Qualitative OD, No (%) | |  |  |  |  |  |  |  |
|  | Parosmia | 26 | (29.9) |  | 8 | (11.9) |  | 0.010* |
|  | Phantosmia | 11 | (12.6) |  | 4 | (6.0) |  | 0.185 |
| Severity of OC obstruction, No (%) | |  |  |  |  |  |  |  |
|  | Absent | 40 | (46.0) |  | 56 | (83.6) |  | <0.000**** |
|  | Mild | 31 | (35.6) |  | 9 | (13.4) |  | 0.003** |
|  | Severe | 16 | (18.4) |  | 2 | (3.0) |  | 0.004** |
| Olfactory tests, median (IQR) | |  |  |  |  |  |  |  |
|  | T&T Olfactometer |  |  |  |  |  |  |  |
|  | Average detection threshold | 1.8 | (1.0-3.6) |  | 4.4 | (3.1-5.6) |  | <0.000**** |
|  | Average recognition threshold | 3.2 | (2.2-4.6) |  | 5.4 | (4.0-5.8) |  | <0.000**** |
|  | OE | 6.0 | (3.3-7.0) |  | 4.0 | (2.0-7.5) |  | 0.066 |
|  | VAS | 30.0 | (8.5-49.0) |  | 8.0 | (2.0-23.0) |  | <0.000**** |
|  | SAOQ | 41.7 | (12.5-74.2) | | 10.3 | (1.9-40.0) |  | 0.002** |
| Retronasal olfactory test (AT), No/n (%) | | |  |  |  |  |  |  |
|  | Prolonged latency | 55/61 | (90.2) |  | 27/35 | (77.1) |  | 0.131 |
|  | Shortened duration | 33/61 | (54.1) |  | 24/34 | (70.6) |  | 0.132 |
|  | No response | 8/70 | (11.4) |  | 27/62 | (43.5) |  | <0.000**** |

†Median age, median disease duration, olfactory tests were analysed with the p-value using the Mann–Whitney U test. The other factors were analysed with p-values using Fisher’s exact test. Statistical significance was set at p < 0.05.

PCOD, post-Covid-19 olfactory dysfunction; PIOD, post-infectious olfactory dysfunction; IQR, interquartile range; OD, olfactory dysfunction; OC, olfactory cleft; SAOQ, Self-administered odour questionnaire; VAS, Visual Analogue Scale for olfactory dysfunction; OE, Open Essence; AT, Alinamin test.

*p < 0.05, **p < 0.01, ****p < 0.000

| Table 2. Demographic characteristics and olfactory test results of PCOD patients according to OC obstruction severity | | | | | | | | | | | | | | | | | |
| --- | --- | --- | --- | --- | --- | --- | --- | --- | --- | --- | --- | --- | --- | --- | --- | --- | --- |
| Severity of OC obstruction | | Absent (n=40) | |  | Mild (n=31) | |  | Severe (n=16) | |  | P  (Unadjusted) |  | Adjusted p-value | | | | |
|  |  |  |  |  |  |  |  |  |  |  |  |  | Absent-Mild |  | Mild-Severe |  | Absent-Severe |
| Sex, No (%) | |  |  |  |  |  |  |  |  |  |  |  |  |  |  |  |  |
|  | Male | 17 | (42.5) |  | 12 | (38.7) |  | 7 | (43.8) |  | 0.928 |  |  |  |  |  |  |
|  | Female | 23 | (57.5) |  | 19 | (61.3) |  | 9 | (56.2) |  |  |  |  |  |  |  |  |
| Age, median yr (IQR) | |  |  |  |  |  |  |  |  |  |  |  |  |  |  |  |  |
|  |  | 41.0 | (28.0-54.0) | | 40.0 | (30.5-55.0) | | 21.0 | (15.8-28.0) | | <0.000**** |  | 0.415 |  | 0.007** |  | <0.000**** |
| Disease duration, median month (IQR) | | |  |  |  |  |  |  |  |  |  |  |  |  |  |  |  |
|  |  | 7.0 | (4.0-12.0) |  | 5.0 | (3.0-8.5) |  | 4.0 | (3.8-5.0) |  | 0.032* |  | 0.180 |  | 0.053 |  | >0.999 |
| Smoking, No (%) | |  |  |  |  |  |  |  |  |  |  |  |  |  |  |  |  |
|  | Current | 3 | (7.5) |  | 1 | (3.2) |  | 0 | (0) |  | 0.433 |  |  |  |  |  |  |
|  | Former or never | 37 | (92.5) |  | 30 | (96.8) |  | 16 | (100) |  |  |  |  |  |  |  |  |
| Qualitative OD, No (%) | |  |  |  |  |  |  |  |  |  |  |  |  |  |  |  |  |
|  | Parosmia | 17 | (42.5) |  | 7 | (22.6) |  | 3 | (18.8) |  | 0.099 |  |  |  |  |  |  |
|  | Phantosmia | 4 | (10.0) |  | 4 | (12.9) |  | 3 | (18.8) |  | 0.672 |  |  |  |  |  |  |
| Olfactory tests, median (IQR) | |  |  |  |  |  |  |  |  |  |  |  |  |  |  |  |  |
|  | T&T Olfactometer |  |  |  |  |  |  |  |  |  |  |  |  |  |  |  |  |
|  | Average detection threshold | 1.4 | (1.0-2.2) |  | 1.8 | (1.0-3.0) |  | 4.3 | (3.6-5.3) |  | 0.001** |  | >0.999 |  | 0.002** |  | 0.001** |
|  | Average recognition threshold | 2.8 | (2.2-4.2) |  | 3.0 | (2.0-4.0) |  | 5.0 | (4.2-5.7) |  | 0.002** |  | >0.999 |  | 0.003** |  | 0.003** |
|  | OE | 6.0 | (5.0-8.0) |  | 6.0 | (3.0-7.0) |  | 2.5 | (0.8-5.5) |  | 0.016* |  | 0.768 |  | 0.191 |  | 0.012* |
|  | VAS | 29.0 | (13.0-48.0) | | 39.0 | (9.0-52.0) |  | 13.5 | (1.8-27.3) |  | 0.039* |  | >0.999 |  | 0.042* |  | 0.091 |
|  | SAOQ | 45.0 | (20.0-77.5) | | 32.4 | (12.5-58.0) | | 6.9 | (0.0-51.4) |  | 0.084 |  |  |  |  |  |  |
| Retronasal olfactory test (AT), No (n) (%) | | |  |  |  |  |  |  |  |  |  |  |  |  |  |  |  |
|  | Prolonged latency | 25 (27) | (92.6) |  | 21 (22) | (95.5) |  | 10 (12) | (83.3) |  | 0.652 |  |  |  |  |  |  |
|  | Shortened duration | 12 (27) | (44.4) |  | 14 (21) | (66.7) |  | 7 (12) | (58.3) |  | 0.386 |  |  |  |  |  |  |
|  | No response | 4 (31) | (12.9) |  | 4 (26) | (15.4) |  | 0 (12) | (0) |  | 0.370 |  |  |  |  |  |  |
| Anosmia, No (n) (%) | |  |  |  |  |  |  |  |  |  |  |  |  |  |  |  |  |
|  | T&T olfactometer | 4 | (10.0) |  | 3 | (9.7) |  | 5 | (31.3) |  | 0.081 |  |  |  |  |  |  |
|  | Both T&T and AT | 4 (31) | (12.9) |  | 2 (26) | (7.7) |  | 0 (12) | (0) |  | 0.393 |  |  |  |  |  |  |

†Median age, median disease duration, olfactory tests were analysed with the p-value using the Kruskal-Wallis test. The other factors were analysed with p-values using Fisher’s exact test. The Dunn-Bonferroni post-hoc test was used to adjust the significance level in multiple comparison tests. Statistical significance was set at p < 0.05.

PCOD, post-Covid-19 olfactory dysfunction; IQR, interquartile range; OD, olfactory dysfunction; OC, olfactory cleft; SAOQ, Self-administered odour questionnaire; VAS, Visual Analogue Scale for olfactory dysfunction; OE, Open Essence; AT, Alinamin test.

* p < 0.05, ** p < 0.01, **** p < 0.000

| Table 3. Demographic characteristics and olfactory test results of PIOD patients according to OC obstruction severity | | | | | | | | | | | |
| --- | --- | --- | --- | --- | --- | --- | --- | --- | --- | --- | --- |
| Severity of OC obstruction | | Absent (n=56) | |  | Mild (n=9) | |  | Severe (n=2) | |  | P  (Unadjusted) |
| Sex, No (%) | |  |  |  |  |  |  |  |  |  |  |
|  | Male | 10 | (17.9) |  | 3 | (33.3) |  | 0 | (0) |  | 0.431 |
|  | Female | 46 | (82.1) |  | 6 | (66.7) |  | 2 | (100) |  |  |
| Age, median yr (IQR) | |  |  |  |  |  |  |  |  |  |  |
|  |  | 64.0 | (54.0-70.3) | | 48.0 | (29.0-67.0) | | 56.5 | (52.3-60.8) | | 0.151 |
| Disease duration, median month (IQR) | | |  |  |  |  |  |  |  |  |  |
|  |  | 5.0 | (3.0-7.0) |  | 5.0 | (2.0-7.0) |  | 3.5 | (3.3-3.8) |  | 0.806 |
| Smoking, No (%) | |  |  |  |  |  |  |  |  |  |  |
|  | Current | 4 | (7.1) |  | 1 | (11.1) |  | 0 | (0) |  | 0.860 |
|  | Former or never | 48 | (85.7) |  | 8 | (88.9) |  | 2 | (100) |  |  |
| Qualitative OD, No (%) | |  |  |  |  |  |  |  |  |  |  |
|  | Parosmia | 8 | (14.3) |  | 0 | (0) |  | 0 | (0) |  | 0.410 |
|  | Phantosmia | 3 | (5.4) |  | 1 | (11.1) |  | 0 | (0) |  | 0.588 |
| Olfactory tests, median (IQR) | |  |  |  |  |  |  |  |  |  |  |
|  | T&T Olfactometer |  |  |  |  |  |  |  |  |  |  |
|  | Average detection threshold | 4.3 | (3.0-5.6) |  | 5.2 | (3.4-5.4) |  | 4.0 | (3.7-4.3) |  | 0.901 |
|  | Average recognition threshold | 5.4 | (4.0-5.8) |  | 5.4 | (4.0-5.6) |  | 5.2 | (5.2-5.2) |  | 0.869 |
|  | OE | 4.0 | (1.0-7.0) |  | 5.0 | (4.0-8.0) |  | 7.0 | (6.0-8.0) |  | 0.271 |
|  | VAS | 7.0 | (1.0-20.5) | | 21.5 | (8.5-42.0) |  | 7.5 | (6.3-8.8) |  | 0.253 |
|  | SAOQ | 10.0 | (0.6-35.2) | | 32.7 | (5.6-87.0) | | 21.5 | (12.5-30.5) |  | 0.528 |
| Retronasal olfactory test (AT), No (n) (%) | | |  |  |  |  |  |  |  |  |  |
|  | Prolonged latency | 23 (28) | (82.1) |  | 5 (5) | (100) |  | 1 (2) | (50) |  | 0.277 |
|  | Shortened duration | 18 (27) | (66.7) |  | 5 (5) | (100) |  | 1 (2) | (50) |  | 0.260 |
|  | No response | 24 (52) | (46.2) |  | 3 (8) | (37.5) |  | 0 (2) | (0) |  | 0.406 |
| Anosmia, No (n) (%) | |  |  |  |  |  |  |  |  |  |  |
|  | T&T olfactometer | 24 | (42.9) |  | 3 | (33.3) |  | 0 | (0) |  | 0.431 |
|  | Both T&T and AT | 19 (52) | (36.5) |  | 2 (8) | (25.0) |  | 0 (2) | (0) |  | 0.479 |

†Median age, median disease duration, olfactory tests were analysed with the p-value using the Kruskal-Wallis test. The other factors were analysed with p-values using Fisher’s exact test. The Dunn-Bonferroni post-hoc test was used to adjust the significance level in multiple comparison tests. Statistical significance was set at p < 0.05.

PCOD, post-Covid-19 olfactory dysfunction; IQR, interquartile range; OD, olfactory dysfunction; OC, olfactory cleft; SAOQ, Self-administered odour questionnaire; VAS, Visual Analogue Scale for olfactory dysfunction; OE, Open Essence; AT, Alinamin test.
